# Depressive symptoms anticipate behavioral and emotional factors among older adults: A prospective cross-lagged panel design

**DOI:** 10.1101/2022.08.31.22279439

**Authors:** Joseph Kazan, Andrew R. Gerlach, Akiko Mizuno, Carmen Andreescu, Howard J. Aizenstein, Scott Ward, Kara J. Buente, Sarah T. Stahl

## Abstract

This study examined the temporal relationship among depression, anxiety, insomnia, perceived stress, and physical activity in adults aged 60+ years with a history of major depressive disorder. We conducted a longitudinal study with 12 weeks of follow-up. Assessments consisted of phone or video interviews and included questionnaires evaluating depression, anxiety, insomnia, perceived stress, and physical activity. Our analytic approach consisted of a depression-focused cross-lagged panel model (CLPM) to examine week-to-week correlations among the five measures. The depression-focused CLPM identified statistically significant week-to-week self-predictive effects for each of the five measures. Higher depressive symptom burden was a strong predictor of increased stress, greater insomnia, and less physical activity the following week. No other cross-measure predictions were statistically significant. Our analytical approach clarifies the directional relationship among variables that typically co-occur with depression showing that higher depression symptom burden predisposes older adults to poor sleep, a reduced level of daytime activity, and a greater sense of stress. These findings support the need for longitudinal assessments and targeted interventions for reducing symptoms of depression in older adults.

## 1. Introduction

As the coronavirus disease 2019 (COVID-19) pandemic spread across the world in early 2020, the World Health Organization began to warn of an accompanying mental health crisis (Nations, 2020). Increase in mortality and morbidity coupled with disruptions in social support systems due to lockdown and physical distancing measures disproportionately affected vulnerable populations in the early stages of the pandemic. Studies conducted between April 2020 and June 2020 in the United States showed a significant increase in symptoms of depression and anxiety when compared with the same period in 2019 (Czeisler et al., 2020).

Despite the wealth of research on the psychological ramifications of the COVID-19 pandemic, very little has been published on the experiences of older adults with depression, a vulnerable group of adults who may have been uniquely impacted by the prolonged stress and physical isolation associated with the pandemic. One study found that older adults with a history of depression experienced worsening depressive symptomatology during the early stages of the pandemic (Oh et al., 2021) presumably due to disruptions in daily routines and lack of access to mental healthcare (Czysz et al., 2021). The increased risk for depression was particularly pronounced among adults aged >70 years and living alone/socially isolated and unable to engage with family and friends (Briggs et al., 2021; Sheffler et al., 2021).

Our study aimed to examine the week-to-week variation among emotional and behavioral factors in older adults with a history of depression. Using cross-lagged panel models, we sought to investigate the directional relationship between depression, anxiety, stress, insomnia, and physical activity. We chose these variables because they frequently co-occur with depression and because of the pandemic’s profound disruption to sleep and physical activity; and increased prevalence of anxiety and stress. Prior research linking depression to emotional and behavioral health has been limited to cross-sectional analyses or longitudinal follow-up over longer time intervals (months to years). This is the first study to examine week-to-week variations in these variables and their relationship over time. This will help determine what constructs are driving the others so that the most efficacious treatment targets can be identified. Given that we conducted this study during the early stages of the COVID-19 pandemic, we also examined the impact of self-reported social isolation on the above-mentioned variables.

## 2. Methods

### 2.1 Study Design

This study included adults aged 60+ years who participated in the Neural Mechanisms of Monoaminergic Engagement in Late-Life Depression Treatment Response study (NEMO R01 MH 076079, NCT03128021) at the University of Pittsburgh. Older adults with major depressive disorder (MDD) received either escitalopram (a selective serotonin reuptake inhibitor) or levomilnacipran (a norepinephrine reuptake inhibitor) for twelve weeks. The goal of the NEMO trial is to understand neurobiological changes associated with successful antidepressant treatment.

Due to the temporary suspension of in-person research activities in the early months of the COVID-19 pandemic, we asked NEMO participants enrolled at that time if they were interested in receiving weekly calls from research staff. From April 2020 to July 2020, we administered a battery of physical and emotional health assessments to 20 participants. Institutional Review Board approval was obtained at the University of Pittsburgh and all participants provided informed consent orally via phone call or video conferencing.

### 2.2 Participants

Participants for NEMO included community dwelling adults aged 60 years and older who met DSM-IV criteria for MDD (using the Structured Clinical Interview for DSM-IV (First and Gibbon, 2004) and reported at least moderate depressive symptom burden defined by a score ≥ 12 on the Montgomery-Asberg Depression Rating Scale (MADRS) (Montgomery and Asberg, 1979). We excluded participants with a history of mania or psychosis, current suicidal ideation, alcohol or substance abuse (current or past 3 months), and dementia of any etiology endorsed via phone screening or diagnosed by Structured Clinical Interview for the Diagnostic and Statistical Manual for Mental Disorders (SCID) (First and Gibbon, 2004). For this secondary analysis, participants who had met at baseline the inclusion criteria for lifetime MDD diagnosis were eligible for weekly phone assessments regardless of current depressive symptomatology (e.g., participants completed NEMO and therefore some experienced a reduction in depression symptoms).

Of the 20 NEMO participants who were interested in receiving weekly calls from research staff, 18 (90%) participants completed two or more phone assessments; three (15%) completed 6 assessments, four (20%) completed 11 timepoints, and eleven (55%) completed all 12 assessments. The 18 participants who completed two or more assessments are the focus of this report.

### 2.3 Measures

Research assessments were conducted over-the phone or by video conference by trained research assistants in geriatric mental health. Demographics were collected at baseline. The emotional and behavioral health questionnaires (detailed below) were collected weekly, for 12 weeks.

#### Demographics

A brief questionnaire assessed basic demographic information including age, sex, race, and years of education.

#### Depression symptoms

Depression symptoms were assessed with the Clinician administered Montgomery-Åsberg Depression Rating Scale (MADRS) (Montgomery and Asberg, 1979). The MADRS assesses the severity of 10 depressive symptoms in patients with mood disorders (Montgomery and Asberg, 1979). Each item is scored on a scale from 0 to 6 for a maximum of 60 points. Higher scores indicate a greater severity of depressive symptomatology where a total score of 0–6 indicates no depression, 7–19 mild depression, 20–34 moderate depression, and >34 severe depression. Depression was analyzed as a continuous variable.

#### Emotional and behavioral health

***Anxiety symptoms*** were assessed with the Hamilton Anxiety Rating Scale (HAM-A), a 14-item clinical clinician-administered assessment of somatic and psychologic symptoms of anxiety (Hamilton, 1959). Items are scored from 0 (not present) to 4 (severe) where a total score of 8-17 indicates mild severity, 18–24 mild to moderate severity and 25–30 moderate to severe. Anxiety was analyzed as a continuous variable. ***Perceived stress*** was self-assessed using the Perceived Stress Scale (PSS) (Cohen et al., 1983). The PSS consists of 10 questions each scored on a scale from 0 (never) to 4 (very often) for a maximum of 40 points. A total score is obtained by reversing responses (e.g., 0 = 4, 1 = 3, 2 = 2, 3 = 1 & 4 = 0) to the four positively stated items (items # 4, 5, 7, and 8) and then summing across all items. Higher scores reflect more severe perceived stress (Cohen et al., 1983). ***Sleep disturbances*** were assessed with the Insomnia Severity Index (ISI) (Bastien et al., 2001) a self-report instrument measuring subjective symptoms of insomnia and the distress associated with the sleep difficulties (Bastien et al., 2001). The 7-items are scored on a scale from 0 to 4 for a maximum of 28 points. Scores greater than 15 indicate clinical insomnia. The ISI was analyzed as a continuous variable. ***Physical activity*** was assessed with the Physical Activity Scale for the Elderly (PASE) (Washburn et al., 1993). The PASE consists of 12 items that measures activities typically chosen by older adults (e.g., walking, leisure activity, housework, and yard work, among others). Each activity is assigned a different weight to account for intensity (e.g., vigorously-intense exercise versus moderately-intense walking). The total score is computed by multiplying the frequency (i.e., yes/no) and the duration (i.e., hours/week) of activity by the item weights and then summing all activities. The total score ranges from 0 to 793, with higher scores indicating greater physical activity (Washburn et al., 1993). ***Social isolation*** was self-assessed with the Patient-Reported Outcomes Measurement Information System, Social Isolation subscale (PROMIS-SI) (2018). This item-bank was developed by the NIH Health Measures and assessed perceptions of isolation from others via a 4-items questionnaire(2018). Each item is scored from 1 to 5 with a maximum score of 20. Higher scores indicate a higher level of self-reported feelings of social isolation.

### 2.4 Statistical Analyses

To examine the associations among emotional and behavioral health, we employed a series of cross-lagged panel models (CLPMs) – a type of structural equation model that describes the relations between observed variables that take the form of variance, covariance, and linear regression. Our model included weekly scores on the MADRS (depression), HAM-A (anxiety), PSS (stress), ISI (insomnia), and PASE (physical activity). All measures were standardized via z-score to allow for scale-invariant comparison. For week-to-week dependence, we tested five CLPMs, each focusing on a single measure acting as a predictor for all other measures the following week *and* an outcome predicted by all other measures the preceding week. For example, in our “depression focused” model, the MADRS at week 2 was treated as a predictor for the MADRS, HAM-A, PSS, ISI, and PASE at week 3 and also an outcome at week 2 predicted by the MADRS, HAM-A, PSS, ISI, and PASE at week 1 (Figure 1). Because all measures were expected to have a reasonable degree of correlation with each other at any given timepoint, we verified that the variance inflation factor was <4 for all variables and included all ten covariances in the model. Each measure was also used as a predictor for itself the following week because we expected strong correlations of each measure over time. This model was repeated with a focus on HAM-A as a predictor and an outcome (model 2), then with PSS (model 3), ISI (model 4), and PASE (model 5) as predictors and outcomes in their respective focused models.

**Figure 1.**
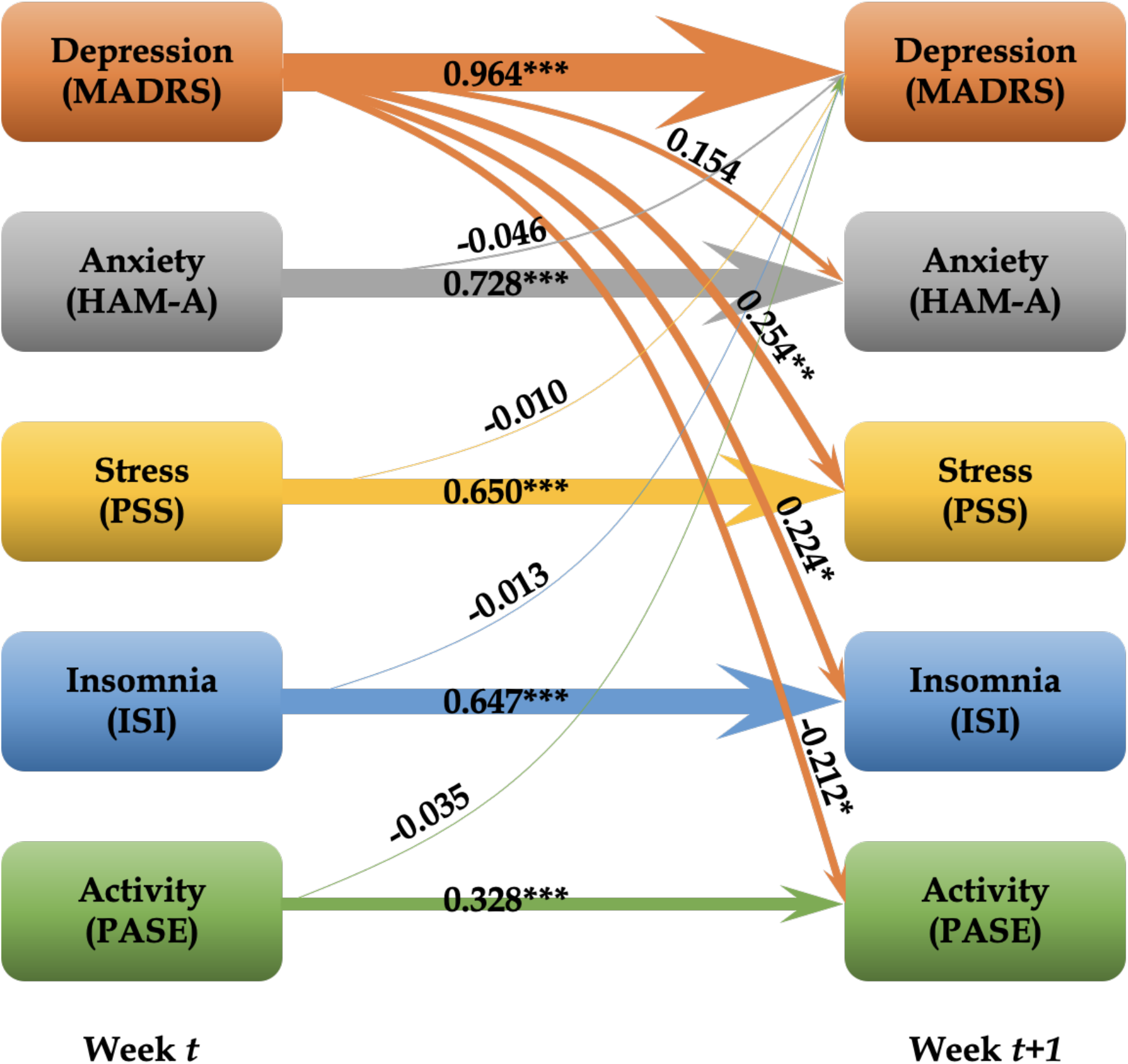
Tested cross-lagged panel model with depression as both a predictor and outcome. Weights of arrows indicate the absolute value of the regression coefficients. Significance levels are indicated by *** p < 0.001, ** 0.001 ≤ p < 0.01, * 0.01 ≤ p < 0.05.

The temporal characteristics of emotional and behavioral health were further examined through smoothing. As shown in Figure 2, an exponential smoothing kernel was applied in both the backward (predictor) and forward (outcome) directions. For a smoothing kernel of size *k*, the predictor (*x*_*t*_) and outcome (*y*_*t*_) values at time *t* are given by:

**Figure 2.**
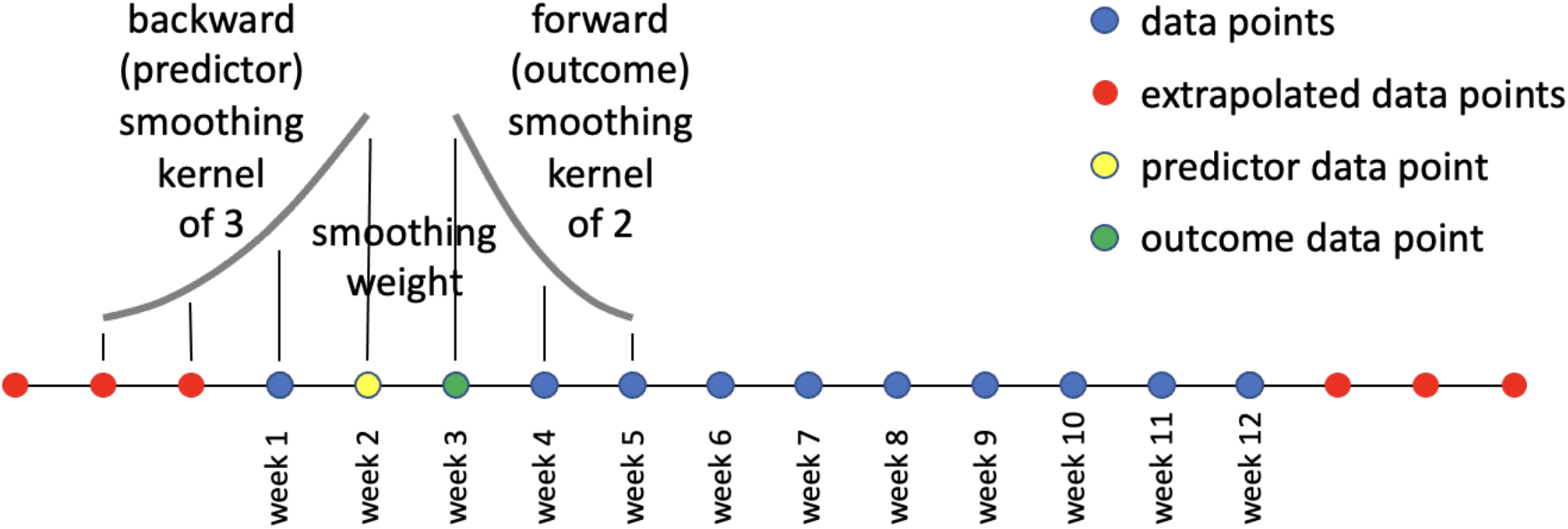
Smoothing of temporal data in the backward (predictor) and forward (outcome) direction. In this example, the backward smoothing kernel is 3 and the forward smoothing kernel is 2, meaning 3 additional previous data points with exponentially decaying weights were used to calculate the predictor value, and a forward smoothing kernel of 2, meaning 2 additional future data points with exponentially decaying weights were used to calculate the outcome value.

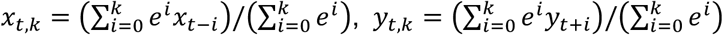

Smoothing kernels from size 0 (no smoothing) to 3 were considered. To maintain a consistent number of data points between the models, we extrapolated data at the beginning and end of the timeline with an initial CLPM estimate as needed. As an additional measure of caution, we compared the reported trends for smoothing to models without extrapolation to qualitatively ensure that the trends were not an effect of extrapolation.

## 3. Results

### 3.1 Descriptive statistics

Sociodemographic characteristics of the study sample are shown in Table 1. Participants’ mean age was 67.75 years (standard deviation [SD] = 6.72 years). Approximately half of the sample were women (55%) and 20% self-reported as ethnic or racial minority individuals. Mean scores on the MADRS (M = 14.35, SD = 9.43) indicated a mild level of depression symptom burden. Participants, on average, reported mild anxiety, moderate stress, subclinical insomnia, and very low levels of physical activity. Compared to participants who completed two or more assessments (n=18) those who declined follow-up assessments (n=2) had slightly higher social isolation scores than participants who remained in the study.

**Table 1.** Participant Characteristics at Baseline (N=20)*

| Characteristic | M (SD) or n (%) |
| --- | --- |
| Age, years | 67.75 (6.72) |
| Female sex, n (%) | 11 (55.0%) |
| White race, n (%) | 16 (78.0%) |
| Education (years) | 15.75 (2.74) |
| Social Isolation (PROMIS-SI)** | 7.00 (4.48) |
| Model variables |  |
| Depression (MADRS) | 14.35 (9.43) |
| Anxiety (HAM-A) | 15.10 (8.91) |
| Stress (PSS) | 18.45 (8.33) |
| Insomnia (ISI) | 9.90 (6.97) |
| Physical activity (PASE) | 153.06 (69.79) |

**Notes.** MADRS = Montgomery-Asberg Depression Rating Scale; HAM-A = Hamilton Anxiety Rating Scale; PSS = Perceived Stress Scale; ISI = Insomnia Severity Index; PASE = Physical Activity Scale for the Elderly
\* 20 participants completed assessments at baseline; participants with more than one time point (n=18) were included in the longitudinal analyses.
\*\* PROMIS scales are computerized adaptive tests with population normed T scores with a mean of 50 and a standard deviation of 10.

### 3.2 Temporal relations among emotional and behavioral health in LLD

Over the 12-week period, all five measures showed general stability: variance was non-zero, but trajectories tended to be flat or nearly so. In each of the five CLPM models (with each focused on one of the studied variables), week-to-week self-predictive effects were statistically significant for all five measures: depression (*P* < .001); anxiety (*P* < .001); stress (*P* < .001); sleep (*P* < .001); and physical activity (*P* < .001). All five variables were predictive of themselves the following week, regardless of the focus of the CLPM model.

In the depression-focused model, depression was also a strong predictor of stress (*β* = 0.254, *P* < .01), insomnia (*β* = 0.224, *P* < .05), and physical activity (*β* = -0.212, *P* < .05) the following week (Table 2). Greater depression at week *t* was associated with more perceived stress, greater insomnia, and less physical activity at week *t+1*. The standardized regression coefficients are shown in Figure 1. In the anxiety-focused model, scores on the HAM-A predicted future HAM-A scores in addition to the PASE (supplemental table 1). Greater anxiety at week *t* was associated with lower physical activity at week *t+1*. In the other three CLPMs (perceived stress, insomnia, and physical activity) each measure at week *t* was predictive of itself at week *t+1* (supplemental tables 2 – 4). No other cross-measure predictions emerged significant.

**Table 2.** Cross-lagged panel model with depression as both a predictor and outcome

| Predictor (*t*) | Outcome (*t*+1) | Beta | Std. Err. | p-value |
| --- | --- | --- | --- | --- |
| Depression | Depression | 0.964 | 0.102 | <.001 |
| Depression | Anxiety | 0.154 | 0.099 | 0.121 |
| Depression | Stress | 0.254 | 0.098 | 0.009 |
| Depression | Insomnia | 0.224 | 0.095 | 0.018 |
| Depression | Physical activity | -0.212 | 0.098 | 0.031 |
| Anxiety | Depression | -0.046 | 0.069 | 0.505 |
| Anxiety | Anxiety | 0.728 | 0.081 | <.001 |
| Stress | Depression | -0.01 | 0.066 | 0.883 |
| Stress | Stress | 0.65 | 0.082 | <.001 |
| Insomnia | Depression | -0.013 | 0.057 | 0.818 |
| Insomnia | Insomnia | 0.647 | 0.076 | <.001 |
| Physical activity | Depression | -0.035 | 0.038 | 0.361 |
| Physical activity | Physical activity | 0.328 | 0.076 | <.001 |

As shown in Figure 2, backward smoothing (including more previous information in the predictor) increased the strength of all five self-predicting regression coefficients in the depression focused CLPM. While the trend is clear, testing for statistical significance was beyond the scope of this analysis. Forward smoothing (including more future information in the outcome) has the opposite effect, generally decreasing the self-predicting regression coefficients, though the effect is far less pronounced than for backward smoothing. However, for the three significant cross-measure regression coefficients (MADRS predicting the following week’s PSS, ISI, and PASE), this trend is reversed for both backward and forward smoothing.

### 3.3 Post-hoc analysis: Social isolation during COVID-19

Because data were collected during the early stages of the COVID-19 pandemic, we explored whether social isolation was associated with emotional and behavioral health. To test this, we performed k-means clustering (MacQueen, 1967) on the week-to-week means of the four PROMIS-Social Isolation questions. Because of the small sample size, we relied on the elbow method to determine the optimal number of clusters. Average scores on the MADRS, HAM-A, PSS, ISI, and PASE were compared between the resulting groups using two sample t-tests. Two distinct groups of high (n=5) and low (n=13) social isolation were identified (see Table 3). Compared to participants with low social isolation, those with high levels of social isolation reported greater depressive symptom burden (*t* = 3.77, *df* = 5.59, *P* = .010); greater anxiety symptom burden (*t* = 4.18, *df* = 15.96, *P* < .001); more perceived stress (*t* = 6.10, *df* = 10.45, *P* < .001); more insomnia symptoms (*t* = 2.68, *df* = 8.31, *P* = .027); and a trend of less physical activity (*t* = -1.88, *df =* 15.81, *P* = .079).

**Table 3.**
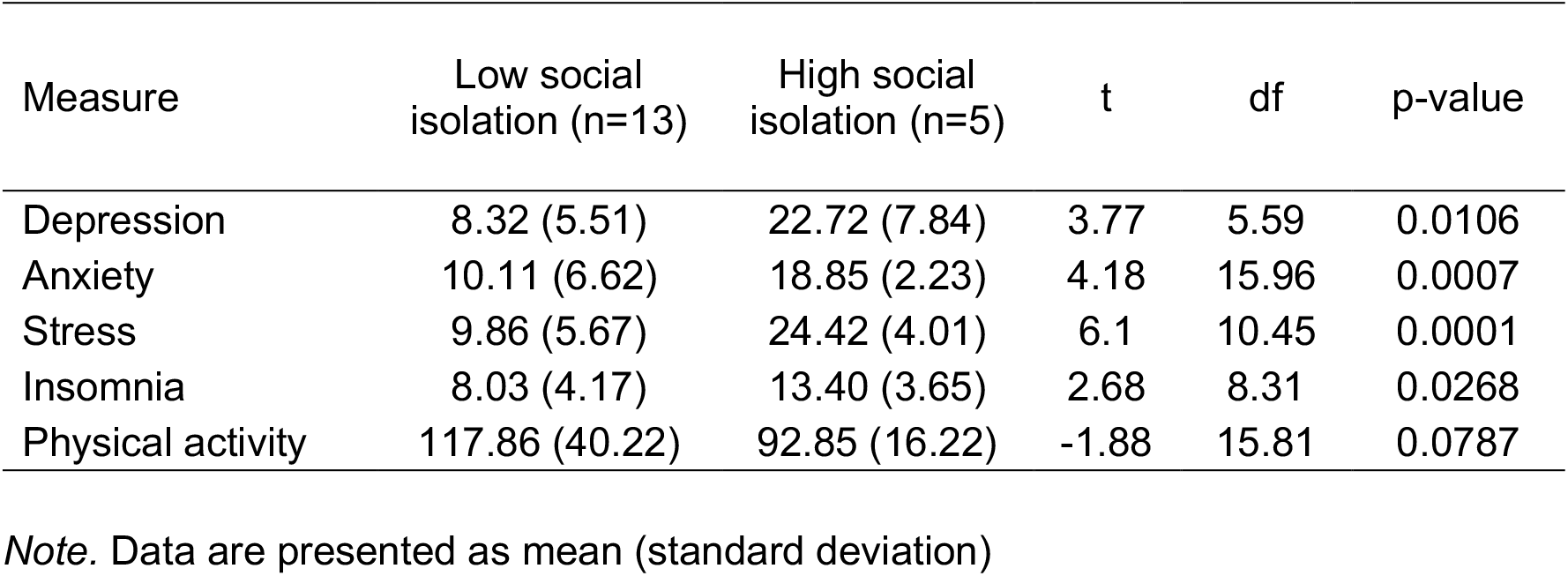
Comparison of depression, anxiety, stress, insomnia, and physical activity measures between low (n=13) and high (n=5) social isolation groups

## 4. Discussion

The longitudinal nature of our analysis and the frequency of time points provided an innovative approach to evaluating the week-to-week directional relationships among depression, anxiety, stress, insomnia, and physical activity. All five variables were significant predictors of themselves the following week, regardless of the focus of the CLPM model. This finding is likely due to the short time interval between assessments. Depression and anxiety are chronic conditions that require frequent monitoring for months to years to achieve meaningful change in symptomatology (Beekman et al., 2002; Lenze et al., 2005). The chronicity of these illnesses could explain why scores on the MADRS and HAM-A are strong predictors of themselves the following week. Depression and anxiety scores typically require a longer follow-up period to observe significant trends (decreasing or increasing) over time.

Our depression-focused CLPM had the most statistically significant predictions when compared to the other four CLPMs. This is not surprising given that our sample consisted of older adults with a history of depression. An increase in depressive symptomatology was follwed by greater insomnia severity, greater perceived stress, and less physical activity the following week. This finding is consistent with many other studies showing that depression co-occurs with insomnia, stress, and physical inactivity (Cristóbal-Narváez et al., 2022; Franzen and Buysse, 2008; Wassink-Vossen et al., 2014). Our analytical approach clarifies the directional relationship among these variables showing that depression predisposes older adults to poor sleep, a reduced level of daytime activity, and a greater sense of stress. From a clinical perspective, these findings suggest that if we aggressively treat and reduce depression, we may be able to prevent its debilitating downstream consequences.

Other studies have shown that perceived stress predicts worsening depression (Cristóbal-Narváez et al., 2022; Rayens and Reed, 2014); we observed the reverse direction, that worsening depression was associated with increased perceived stress the following week. Older adults with a history of depression may have been particularly vulnerable to the negative health effects of COVID-19 which presumably increases feelings or thoughts regarding the amount of stress they were experiencing during the early stages of pandemic. Additionally, the deleterious effects of perceived stress on inflammation and cardiovascular health are well documented (Jain et al., 2007; Strine et al., 2004). Studies have shown that exposure to socio-environmental stress is linked to a pro-inflammatory state (Slavich and Irwin, 2014). This phenomenon is in turn associated with both the pathogenesis of depression as well as several somatic conditions including cardiovascular diseases (Cristóbal-Narváez et al., 2022; Slavich and Irwin, 2014). It is possible that older adults with a history of depression are at high risk for other physical, behavioral, and emotional consequences associated with the increased stress experienced during COVID-19.

In terms of insomnia, many studies show that depression is the most common cause of insomnia in older adults (Buysse et al., 1994; Ohayon et al., 1998). Similarly, it has been estimated that 90% of patients with depression also experience sleep disturbances (Tsuno et al., 2005). Severity of depression is further associated with the severity of insomnia and late-life depression may also be more difficult to treat in the presence of insomnia (Dew et al., 1997). Our findings show that an increase in depressive symptomatology is predictive of worse insomnia the following week. Given the overlap in symptomatology, several behavioral interventions such as cognitive behavioral therapy for insomnia (CBTI) have previously been successful in improving co-occurring depression (Franzen and Buysse, 2008). Our results re-affirm the importance of developing comprehensive strategies that treat depression and insomnia in tandem among older adults.

Depressive symptomatology was also predictive of less physical activity the following week. The physical activity scale for the elderly (PASE) score included both occupational activity and leisure activity. Numerous studies have shown that late-life depression is associated with lower levels of physical activity(Lindwall et al., 2011; Wassink-Vossen et al., 2014). These results are relevant given that physical inactivity is considered by the World Health Organization to be a major risk factor for mortality in the world (Organization, 2004). A lifestyle with persistently low physical activity is associated with most chronic diseases as well as the ten leading causes of death in the United States(Booth et al., 2017; Organization, 2004). Consequently, researchers in healthy aging have extensively studied physical exercise interventions in the management of depression in older adults (Aguiñaga et al., 2018; Kok and Reynolds, 2017). A recent meta-analysis showed that physical exercise interventions resulted in a reduction in depressive symptoms in older adults (Klil-Drori et al., 2020). Our results highlight the reciprocal relationship between increased physical activity and improvement in depressive symptomatology in the context of healthy aging.

In our post-hoc analysis, participants who perceived themselves as more socially isolated reported greater depressive symptom burden, greater anxiety, more perceived stress, and worse insomnia. These findings are supported in the literature, with published data overwhelmingly supporting the deleterious effects of loneliness and social isolation on the mental and physical health of older adults (Cornwell and Waite, 2009; Hawkley et al., 2010; Kotwal et al., 2021). This is particularly relevant given that older adults reported increased feelings of loneliness and perceived isolation during the COVID-19 pandemic (Emerson, 2020; Kotwal et al., 2021). These feelings were also associated with worsening depression and anxiety symptoms, which re-affirms the need for a holistic approach to target both social and behavioral components of geriatric mental health (Kotwal et al., 2021).

Several limitations of this study should be acknowledged. First, our assessment of social connectedness addressed functional connectedness only (e.g., perceptions of social support and loneliness) and did not include structural (social isolation) or quality-related dimensions (e.g., positive and negative aspects) of social connectedness(Lutz et al., 2021). Measurement approaches that include several dimensions of social relationships will help identify mechanisms and treatment targets for improving social connectedness among depressed older adults. Second, we conducted this study during the early stages of the COVID-19 pandemic when older adults were extremely vulnerable to the health effects of social isolation due to government-mandated stay-at-home orders and physical distancing. Our findings may highlight “extreme” social isolation and should be replicated in larger samples of older adults and/or compared to social isolation data collected in later stages of the COVID-19 pandemic. Third, our sample included a disproportionate number of White participants compared to the number of participants who identify as racial or ethnic minority individuals. Given growing evidence of the differential impact of the COVID-19 pandemic on various racial and ethnic minority groups, more research is needed to diversify our understanding of the relevant issues affecting older adults’ mental health. Fourth, our modest sample size limits our ability to analyze subtler relationships in the data. However, this is somewhat ameliorated by the repeated measures within subjects. Finally, the variables tested (insomnia, stress, anxiety, activity) may all be symptoms of the depressive syndrome. The downstream consequences reported here may be manifestations of same illness from one week to the next.

In conclusion, we found evidence from a sample of older adults with a history of depression, that depression symptoms play an influential role in the onset of other emotional and behavioral health factors including greater insomnia burden, increased stress, and physical inactivity. Strengths of this study include the use of panel data with multiple observations from each participant over time. This innovative method allowed for the analyses of dynamic systems to describe the oscillation of emotional and behavioral health over a 12-week period. Translated into practice, our findings suggest that if we target and reduce depression symptoms in older adults, we may be able to prevent or delay the onset of its sequelae.

## Data Availability

All data produced in the present study are available upon reasonable request to the authors.

## Description of author’s roles

JK made substantial contributions to the conception or the design of the study and the drafting and revising of the manuscript. AG made substantial contributions to the acquisition, analysis, and interpretation of data for the work, and the writing and revising of the manuscript. SW and KJB were involved with the acquistion of data and manuscript preparation. AM, CA, HJA, and STS were involved with conception and design of the study, data interpretation, and manuscript preparation. All authors approved the final version of this manuscript.

