## Supplemental Tables for "Depressive symptoms anticipate behavioral and emotional factors among older adults: A prospective cross-lagged panel design"

*Supplemental Table 1. Cross-lagged panel model with anxiety as both a predictor and outcome*

| Predictor (*t*) | Outcome (*t+1*) | Beta | Std. Err. | p-value |
| --- | --- | --- | --- | --- |
| Depression | Depression | 0.768 | 0.074 | <.001 |
| Depression | Anxiety | -0.072 | 0.098 | 0.463 |
| Anxiety | Depression | 0.11 | 0.087 | 0.206 |
| Anxiety | Anxiety | 0.852 | 0.098 | <.001 |
| Anxiety | Stress | 0.166 | 0.085 | 0.05 |
| Anxiety | Insomnia | 0.16 | 0.088 | 0.068 |
| Anxiety | Physical Activity | -0.188 | 0.093 | 0.043 |
| Stress | Anxiety | 0.062 | 0.072 | 0.387 |
| Stress | Stress | 0.71 | 0.069 | <.001 |
| Insomnia | Anxiety | 0.025 | 0.067 | 0.712 |
| Insomnia | Insomnia | 0.681 | 0.073 | <.001 |
| Physical Activity | Anxiety | -0.011 | 0.045 | 0.807 |
| Physical Activity | Physical Activity | 0.324 | 0.076 | <.001 |

*Supplemental Table 2. Cross-lagged panel model with stress as both a predictor and outcome*

| Predictor (*t*) | Outcome (*t+1*) | Beta | Std. Err. | p-value |
| --- | --- | --- | --- | --- |
| Depression | Depression | 0.808 | 0.072 | <.001 |
| Depression | Stress | 0.144 | 0.104 | 0.166 |
| Anxiety | Anxiety | 0.738 | 0.066 | <.001 |
| Anxiety | Stress | 0.048 | 0.081 | 0.549 |
| Stress | Depression | 0.092 | 0.086 | 0.283 |
| Stress | Anxiety | 0.14 | 0.084 | 0.098 |
| Stress | Stress | 0.732 | 0.096 | <.001 |
| Stress | Insomnia | 0.135 | 0.085 | 0.112 |
| Stress | Physical Activity | -0.17 | 0.092 | 0.064 |
| Insomnia | Stress | -0.029 | 0.07 | 0.672 |
| Insomnia | Insomnia | 0.706 | 0.069 | <.001 |
| Physical Activity | Stress | 0.011 | 0.048 | 0.825 |
| Physical Activity | Physical Activity | 0.35 | 0.074 | <.001 |

*Supplemental Table 3. Cross-lagged panel model with insomnia as both a predictor and outcome*

| Predictor (*t*) | Outcome (*t+1*) | Beta | Std. Err. | p-value |
| --- | --- | --- | --- | --- |
| Depression | Depression | 0.794 | 0.064 | <.001 |
| Depression | Insomnia | 0.112 | 0.107 | 0.297 |
| Anxiety | Anxiety | 0.752 | 0.067 | <.001 |
| Anxiety | Insomnia | 0.047 | 0.089 | 0.596 |
| Stress | Stress | 0.758 | 0.067 | <.001 |
| Stress | Insomnia | -0.022 | 0.082 | 0.786 |
| Insomnia | Depression | 0.069 | 0.079 | 0.383 |
| Insomnia | Anxiety | 0.094 | 0.083 | 0.256 |
| Insomnia | Stress | 0.082 | 0.081 | 0.312 |
| Insomnia | Insomnia | 0.713 | 0.092 | <.001 |
| Insomnia | Physical Activity | -0.194 | 0.09 | 0.032 |
| Physical Activity | Insomnia | 0.05 | 0.052 | 0.336 |
| Physical Activity | Physical Activity | 0.338 | 0.074 | <.001 |

*Supplemental Table 4. Cross-lagged panel model with physical activity as both a predictor and outcome*

| Predictor (*t*) | Outcome (*t+1*) | Beta | Std. Err. | p-value |
| --- | --- | --- | --- | --- |
| Depression | Depression | 0.777 | 0.054 | <.001 |
| Depression | Physical Activity | -0.094 | 0.152 | 0.538 |
| Anxiety | Anxiety | 0.761 | 0.058 | <.001 |
| Anxiety | Physical Activity | -0.054 | 0.127 | 0.669 |
| Stress | Stress | 0.757 | 0.06 | <.001 |
| Stress | Physical Activity | 0.015 | 0.119 | 0.9 |
| Insomnia | Insomnia | 0.736 | 0.062 | <.001 |
| Insomnia | Physical Activity | -0.087 | 0.11 | 0.43 |
| Physical Activity | Depression | -0.085 | 0.061 | 0.166 |
| Physical Activity | Anxiety | -0.063 | 0.065 | 0.327 |
| Physical Activity | Stress | -0.077 | 0.064 | 0.231 |
| Physical Activity | Insomnia | -0.031 | 0.066 | 0.639 |
| Physical Activity | Physical Activity | 0.336 | 0.079 | <.001 |
